# Association of race, ethnicity, and area deprivation with the prevalence of atrial fibrillation in a large US population

**DOI:** 10.1101/2025.06.24.25330201

**Authors:** Alvaro Alonso, Gabriel Najarro, Amit J. Shah, Linzi Li, Tené T. Lewis

**Affiliations:** Department of Epidemiology, Rollins School of Public Health, Emory University, Atlanta, GA; Division of Cardiology, Department of Medicine, Emory University School of Medicine, Atlanta, GA

## Abstract

**Objective:** To examine the association of atrial fibrillation (AF) prevalence with neighborhood deprivation in a large, national U.S. population across racial and ethnic groups and its association.

**Methods:** We analyzed data from Epic Cosmos, comprising over 56 million adults with demographic and area deprivation index data in 2024. AF prevalence was defined using ICD-10 codes.

**Results:** Overall AF prevalence was 3.7%, higher in men and older adults. Prevalence varied by race/ethnicity, highest in White and American Indian/Alaska Native individuals, and lowest in Asian and Hispanic individuals. AF prevalence increased with higher deprivation across most groups.

**Conclusion:** Area-level socioeconomic deprivation was independently associated with higher AF prevalence across all racial and ethnic groups. These findings underscore the importance of addressing structural determinants of health to reduce the burden of AF in diverse communities.

---

Atrial fibrillation (AF) is a common cardiac arrhythmia and an established risk factor for stroke and heart failure. Studies consistently report higher AF burden in White individuals compared to other racial groups, despite a lower burden of risk factors. This paradox remains unexplained, though one hypothesis is underascertainment of AF in minoritized groups due to their higher likelihood of residence in impoverished communities, which might limit access to adequate healthcare and AF diagnosis.^1^ To date, no prior studies have simultaneously examined the association of race, ethnicity, and neighborhood socioeconomic status with AF incidence in the US.

To examine AF prevalence by race, sex, and community-level socioeconomic status, we analyzed data from Epic Cosmos, which includes electronic health records (EHRs) from patients receiving care from healthcare organizations (>1,700 hospitals, >40,000 clinics) using Epic software.^2^ We utilized Epic SlicerDicer, a self-service tool that enables users to query and slice de-identified patient information across multiple dimensions. We calculated AF prevalence, defined as a charge-associated billing diagnoses with an International Classification of Disease 10^th^ edition Clinical Modification I48.x, by sex, race, and national area deprivation index (ADI).^3^ We included base patients (at least two qualifying encounters within a two-year period) ≥18 years in Epic Cosmos during 2024. Race and ethnicity were obtained from EHR records. Individuals of Hispanic ethnicity were categorized as Hispanic, regardless of race. Non-Hispanic individuals were classified as White, Black, Asian, American Indian/Alaska Native, Native Hawaiian/Other Pacific Islander, or Other Race. Prevalence was age-standardized using the 2024 Epic Cosmos age distribution as the reference. Adjusted prevalence ratios (PR) and 95% confidence intervals (CI) were calculated using log-binomial regression. This study was exempt from institutional review board approval and the need for informed consent because it includes deidentified data.

Epic Cosmos included 108,020,223 base patients ≥18 years in 2024, of whom 54,487,100 (50%) had demographic and ADI information. Among these, 58% were female and 42% male, and 47% were <50 years old, 39% were 50-<75 years old, and 14% were ≥75 years old. Race and ethnicity distribution was 65% White, 13% Black, 12% Hispanic, 6% Other Race, 3% Asian, 1% American Indian/Alaska Native and <1% Native Hawaiian/Other Pacific Islanders. Overall AF prevalence was 3.7%. Age-standardized prevalence was higher in males (4.8%) than females (2.8%) and increased with age: 0.3% in individuals aged 18-<50 years, 4.0% in those 50-<75 years, and 14.2% in those ≥75 years. Age-standardized AF prevalence also varied by race and ethnicity: 4.0% White, 2.7% Black, 2.0% Hispanic, 3.7% Other Race, 1.8% Asian, 3.6% American Indian/Alaska Native, and 3.2% Native Hawaiian/Other Pacific Islanders. The **Figure** presents the race and ethnicity-specific age-standardized AF prevalence across ADI deciles stratified by sex. For most groups, higher ADI was associated with increased AF prevalence. For example, one decile increase in ADI was associated with a 3% increase in AF prevalence in White individuals (PR 1.028, 95%CI 1.027-1.028) and a 4% increase in Black individuals (PR 1.042, 95%CI 1.040-1.044). Differences by race and ethnicity were consistent across ADI deciles, with the highest prevalence in White, Other Race, and American Indian/Alaska Native individuals, intermediate prevalence in Black individuals, and the lowest prevalence in Asian and Hispanic individuals. Trends in Native Hawaiian/Other Pacific Islanders were inconsistent, likely due to small sample size. Similar patterns were observed for males and females.

**FIGURE.**
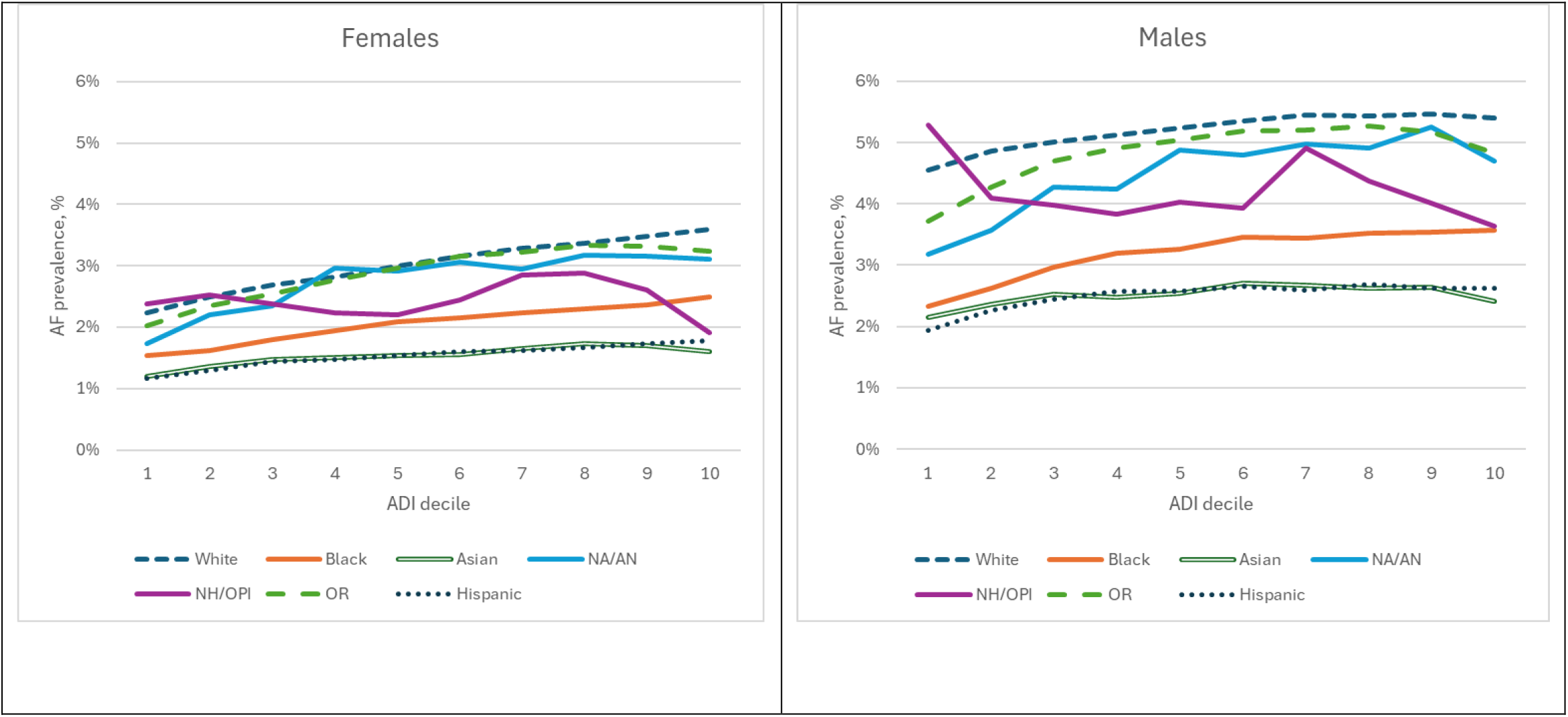
Age-standardized prevalence of atrial fibrillation by race and Area Deprivation Index (ADI), stratified by sex, EPIC Cosmos 2024. NA/AN: Native American/Alaska Native; NH/OPI: Native Hawaiian/Other Pacific Islander; OR: Other race.

Our results align with findings for other cardiovascular diseases, showing higher AF prevalence among individuals in more deprived neighborhoods. Although ADI is just a proxy for neighborhood socioeconomic status and does not capture individual socioeconomic status or access to quality healthcare, our findings do not support the hypothesis that underdiagnosis due to limited care access resulting from community-level disenfranchisement explains the racial paradox in AF burden.

Alternative explanations for the AF paradox include ancestry-specific genetic factors correlated with race, which may contribute to higher AF rates among individuals of European descent.^4^ Alternatively, competing risks, such as higher early mortality in Black individuals, may reduce the number of individuals susceptible to AF at older ages, resulting in the observed pattern.^1^ Further research is needed to evaluate these hypotheses.

This study has limitations, including reliance on EHR data for diagnoses, inclusion only of individuals with some degree of healthcare access, misclassification of race and ethnicity in EHR records, and lack of individual-level comorbidity data, precluding adjustment for clinical variables. Strengths include the large sample size, the demographic similarity of the Epic Cosmos to the US population, and the use of ADI as a validated measure of area-level socioeconomic status.

In summary, we observed that individuals living in more deprived areas had higher AF prevalence and that racial differences in AF prevalence previously reported were consistent across all deprivation levels. Alleviating area-level socioeconomic disparities and uncovering the biological and social mechanisms driving these patterns are critical steps toward reducing the burden of this common arrhythmia.^5^

## Data Availability

All data produced are available online at Epic Cosmos

## Notes

### Competing Interest Statement

The authors have declared no competing interest.

### Funding Statement

This study did not receive any funding.

### Author Declarations

Ethics committee/IRB of Emory University waived ethical approval for this work

## REFERENCES

1. Essien UR, Kornej J, Johnson AE, Schulson LB, Benjamin EJ, Magnani JW. Social determinants of atrial fibrillation. Nat Rev Cardiol. 2021;18:763–773.

2. Tarabichi Y, Frees A, Honeywell S, Huang C, Naidech AM, Moore JH, Kaelber DC. The Cosmos Collaborative: A Vendor-Facilitated Electronic Health Record Data Aggregation Platform. ACI open. 2021;5:e36–e46.

3. Kind AJH, Buckingham WR. Making Neighborhood-Disadvantage Metrics Accessible - The Neighborhood Atlas. N Engl J Med. 2018;378:2456–2458.

4. Marcus GM, Alonso A, Peralta CA, Lettre G, Vittinghoff E, Lubitz SA, Fox ER, Levitzky YS, Mehra R, Kerr KF, et al. European ancestry as a risk factor for atrial fibrillation in African Americans. Circulation. 2010;122:2009–2015.

5. Benjamin EJ, Thomas KL, Go AS, Desvigne-Nickens P, Albert CM, Alonso A, Chamberlain AM, Essien UR, Hernandez I, Hills MT, et al. Transforming Atrial Fibrillation Research to Integrate Social Determinants of Health: A National Heart, Lung, and Blood Institute Workshop Report. JAMA Cardiol. 2023;8:182–191.

